# Dense Breast Tissue Segmentation under Limited Annotation: Comparing CNNs, Transformers, Self-Supervision, and Prompting

**DOI:** 10.64898/2026.02.23.26346855

**Authors:** Bowen Qu, Weixin Liu, Li Zhou, Xingyi Guo, Bradley Malin, Zhijun Yin

## Abstract

Dense breast tissue reduces the sensitivity of mammographic screening and is an important risk factor for breast cancer, making accurate dense-tissue segmentation particularly important when expert pixel-level annotations are limited. We empirically study dense breast tissue segmentation under limited pixel-level supervision using a small expert-annotated dataset (596 images) together with an in-domain unlabeled mammography dataset (20,000 images). We compare classical pixel-level segmentation methods, CNN and U-Net encoder–decoder models, transformer-based models, self-supervised pre-training, fine-tuning strategies, loss functions, and prompt-based MedSAM variants under a unified evaluation protocol. CNN-based models achieve the best standalone segmentation performance while providing the most favorable balance between accuracy and computational cost. In contrast, the adaptive fuzzy c-means + SVM baseline and transformer-based models perform worse in this low-label setting. Our analysis of prompt-based MedSAM further shows that SAM-style models are highly sensitive to prompt quality. Box and breast-region prompts lead to poor performance, whereas CNN-generated dense prompts substantially improve MedSAM performance. However, even with mask-decoder fine-tuning, CNN-prompted MedSAM does not outperform the best standalone CNN model, suggesting that current SAM-style models require stronger prompts or adaptation beyond the mask decoder for diffuse breast-density segmentation. Additionally, a density-calibrated hybrid loss primarily improves percentdensity calibration by reducing the test MAE from 14.8 to 11.8 percentage points. Our computational cost analysis indicates that many generic self-supervised learning and parameter-efficient adaptation strategies provide limited practical benefit relative to their additional cost. These findings offer practical guidance for developing reproducible dense breast tissue segmentation models in mammography studies with limited labeled data.

## I. Introduction

Breast cancer is the most commonly diagnosed cancer among women, with approximately one in eight women in the United States expected to develop invasive disease during their lifetime [1]. Mammography is the standard examination for early detection, but its accuracy decreases in women with high mammographic density, the proportion of radiopaque fibroglandular tissue visible in the image. Dense tissue is both a strong breast cancer risk factor [2] and a structure that can obscure tumors, reducing screening sensitivity [3], [4]. Accurate dense-tissue segmentation is therefore important for quantitative risk assessment, percent-density estimation, and downstream clinical image-analysis workflows. Because percent density is the ratio of dense-tissue area to breast area, systematic over- or under-segmentation biases density estimates even when overlap metrics are high.

Prior work on automated breast density assessment has largely followed two directions. One predicts Breast Imaging Reporting and Data System (BI-RADS) density categories, which assign mammograms to ordinal classes A through D based on radiologist interpretation [5], [6]. These labels are clinically useful but too coarse for applications requiring continuous density measurements, including genetic studies and multi-cohort meta-analyses [7], [8]. The other produces spatially explicit density maps through pixel-level segmentation, including multitask models that jointly segment the breast boundary and dense regions [9] and Deep-LIBRA, a pipeline for validated spatial density maps in risk assessment [10].

Many high-performing systems rely on large private clinical datasets that cannot be released due to regulatory and privacy constraints. Although source code may be available, trained weights and expert annotations are often inaccessible. Reproducing or extending these methods therefore requires large annotated cohorts and substantial computation. Public datasets with expert pixel-level dense-tissue labels remain small and heterogeneous. Even recent datasets with breast-area and dense-tissue masks [11] are modest relative to modern deep learning requirements. More broadly, reliable pixel-level annotation in medical imaging requires specialized clinical expertise and considerable labeling time [12], motivating methods that remain reliable under limited supervision.

Self-supervised learning (SSL), transformer-based vision models, and medical foundation models may reduce dependence on large labeled datasets [13], [14], but they raise practical questions. SSL can incur substantial pre-training cost, and generic pretext tasks may not transfer well to diffuse, low-contrast dense tissue. Transformer models may lack the local inductive bias needed for precise boundaries in small medical datasets. Segment Anything Model (SAM)-style foundation models also depend strongly on prompt quality, making evaluation sensitive to whether prompts are coarse boxes, breast-region masks, or model-generated dense predictions. A fair comparison should therefore evaluate accuracy together with prompt design, fine-tuning, percent-density calibration, and computational cost.

We empirically study dense breast tissue segmentation and percent-density estimation under limited pixel-level supervision. Our two-stage pipeline first extracts the breast region and then segments dense tissue, with optional prompt-based MedSAM refinement. Under a unified protocol, the study includes adaptive fuzzy c-means with SVM, CNN/U-Net models, ViT/DINOv3, and MedSAM-style models; a prompt-aware MedSAM analysis with box-only, box + breast-region, and CNN-derived dense prompts; in-domain SSL and fine-tuning studies, including multi-view SimCLR pre-training and full, layer-wise, and parameter-efficient adaptation; and density-calibrated loss and compute-cost analyses for practical accuracy–efficiency trade-offs. Our main findings are that (i) compact CNN encoder–decoders outperform transformer- and SAM-style alternatives in this limited annotation setting; (ii) generic SSL and parameter-efficient adaptation give little or no gain over ImageNet initialization with full fine-tuning; (iii) MedSAM with a fine-tuned mask decoder does not exceed the CNN prediction used as its prompt; and (iv) a densitybias term mainly improves percent-density calibration rather than mask overlap. Code, data splits, and trained weights will be released to ensure reproducibility.

## II. Methods

### A. Segmentation Pipeline

We use a two-stage mammography pipeline (Figure 1). A breast-region segmentor first generates a binary breast mask from the raw mammogram, removing background and pectoral regions. The masked image is then passed to a dense-tissue segmentor that predicts a binary map separating fibroglandular from fatty tissue. The breast-region segmentor was trained separately using breast-area masks from the Mammogram Density Assessment dataset, with the same patient-level training and validation split used downstream. We selected the checkpoint with the highest validation Dice and froze it for all second-stage experiments. Test-set dense-tissue labels were not used to train or tune this model. Due to the page limit and our focus on dense-tissue segmentation rather than breast-boundary extraction, we treat the first-stage model as fixed preprocessing. All learned models are evaluated in the second stage under the same preprocessing and evaluation protocol.

**Fig 1:**
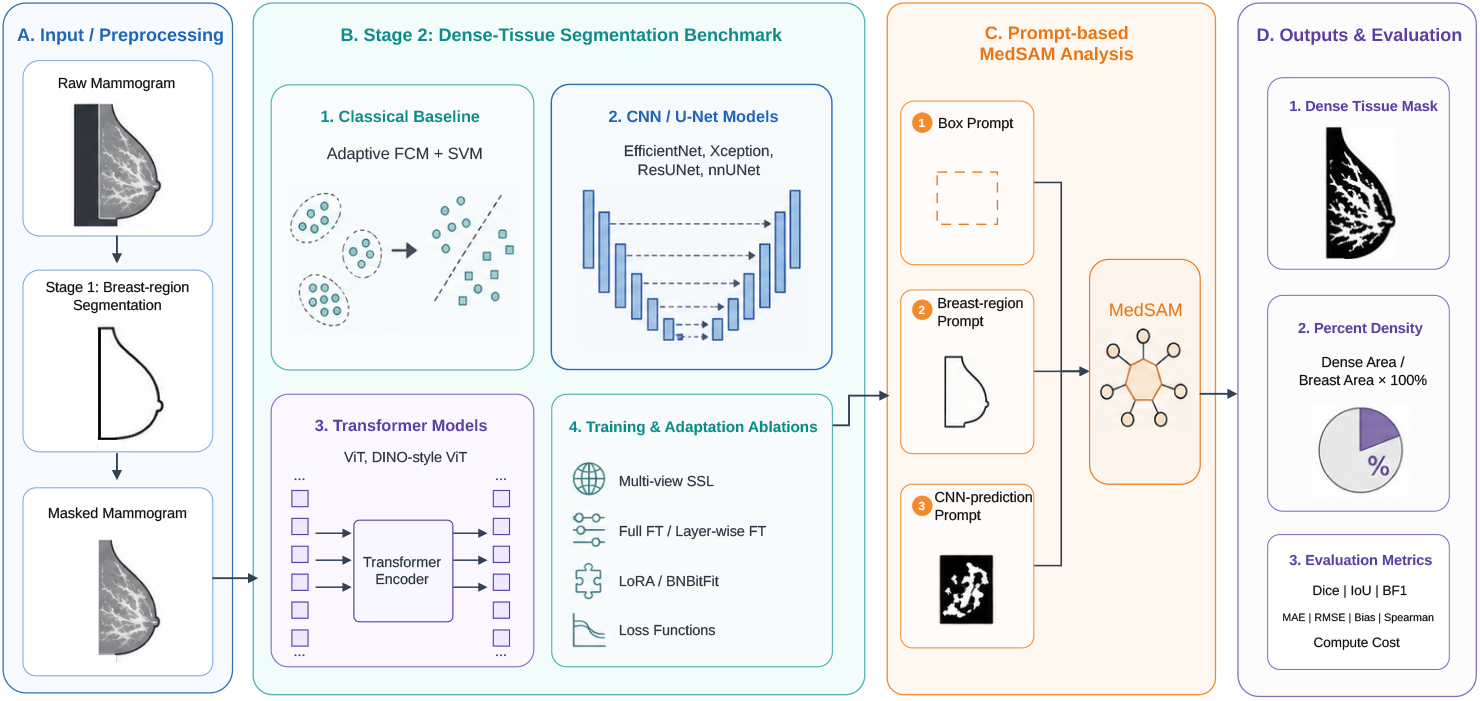
Overview of the evaluation framework. A fixed breast-region model is followed by dense-tissue segmentation, optional MedSAM refinement with box, breast-region, or CNN-prediction prompts, and evaluation of masks, percent density, segmentation quality, calibration, and computational cost.

### B. Percent-Density Estimation and Calibration

Beyond pixel-level overlap, we assess each model by the percent density it implies. For a binarized dense-tissue prediction *ŷ*_*i*_, reference label *y*_*i*_, and breast-region mask *m*_*i*_, predicted and reference percent densities are

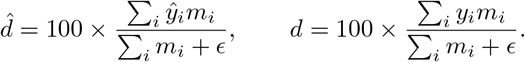

We quantify percent-density calibration by *mean absolute error (MAE), root mean squared error (RMSE)*, and *bias*, defined as the mean of 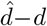 in percentage points, together with Spearman correlations. For external datasets without pixel-level labels, we report the Spearman correlation between 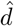and the available clinical BI-RADS density categories.

### C. Classical Pixel-Level Baseline

We include adaptive fuzzy c-means clustering with support vector machine (SVM) classification [15] as a non-deep-learning baseline. The breast-masked mammogram is clustered by tissue intensity, and pixel- or region-level intensity features are passed to an SVM to separate dense from non-dense tissue. The resulting mask is evaluated with the same segmentation and density-estimation metrics as the learned models.

### D. Model Architectures

We evaluate classical pixel-level methods, convolutional encoder–decoder models, transformer-based models, and prompt-based foundation models under the same dense-tissue segmentation protocol unless otherwise stated. VGG19 uses stacked 3 × 3 convolutions and pooling [16]. ResNet-50 adds identity skip connections for deeper optimization [17]. Xception uses depthwise–separable convolutions to decouple spatial and channel mixing [18]. EfficientNet jointly scales depth, width, and resolution for accuracy–efficiency trade-offs; we use EfficientNet-B0 [19]. These CNN encoders are paired with a U-Net-style decoder.

We also evaluate vanilla U-Net [20], ResUNet [21], and nnU-Net-style configurations [22] as task-specific encoder– decoder baselines. Vision Transformer (ViT-Base) uses patch tokens and self-attention for global context [23]. DINOv3 denotes a ViT-style encoder initialized with large-scale self-supervised DINOv3 pre-training [24]. MedSAM [25] is evaluated as a prompt-based medical foundation model rather than a direct replacement for the CNN head; its prompt settings are described in Section II-E.

### E. Prompt-Based MedSAM Refinement

We evaluate MedSAM with three prompts: a coarse box, the box together with a breast-area prompt from the first-stage mask, and a dense prompt from the best standalone CNN without a box. The box is the bounding box of the annotated breast area. Dense prompts are supplied as mask prompts. For each prompt setting, a separate mask decoder is fine-tuned on the training split while the image encoder and prompt encoder are kept frozen; in the CNN-prediction setting, the decoder is trained with CNN predictions on training images, and the prompting CNN is trained only on the training split. The CNN-prediction setting represents a deployable refinement scenario. This analysis tests whether MedSAM refinement can improve over the CNN prediction it receives.

### F. Self-Supervised Learning Strategies

Continuous pretraining through SSL with domain-specific data may further improve model performance [26], [27]. In this task, we continuously pre-train encoders with four popular self-supervised objectives using in-domain, unlabeled data and transfer them to downstream dense tissue segmentation.

SimCLR increases agreement between two stochastic augmentations of the same image while contrasting them against other batch views [28]. Given a mini-batch of *N* images with two augmentations per image (2*N* views), let *z*_*i*_ be the *l*_2_-normalized embedding of view *i, τ* the temperature, and sim(·,·) cosine similarity. For anchor *i*, the positive partner *p*(*i*) is the other augmented view of the same image. The InfoNCE loss is

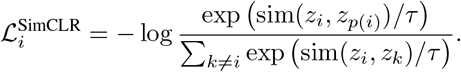

The batch loss averages 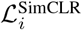 over all anchors.

Multi-view SimCLR extends contrastive learning to four-view mammography, *V* = {L-CC, R-CC, L-MLO, R-MLO} . For anchor *i* and positive view *p* ∈ *P*(*i*), weights *w*_*ip*_ encode anatomical similarity across side and view. A batch of *N* subjects with two augmentations per view yields 8*N* embeddings, with views from other subjects serving as negatives. The multipositive weighted InfoNCE loss [29] is

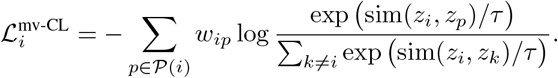

This keeps the standard contrastive denominator while using four-view anatomy to define multiple positives with graded similarity.

Barlow Twins aligns two augmented views without explicit negatives by driving the feature cross-correlation toward the identity [30]. For normalized embeddings *Z*_*a*_, *Z*_*b*_ ∈ ℝ^*B*×*d*^ in a batch of size *B*, let 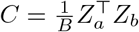 . The objective is

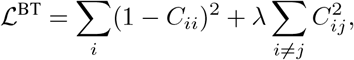

which encourages invariance on the diagonal and reduces off-diagonal redundancy.

Masked image modeling (MIM) masks random patches and reconstructs the missing content, following masked autoencoding [31]. Let *M* be the masked patch set, *x*_*p*_ the ground-truth patch signal, and 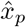 the prediction from an encoder–decoder that receives the masked image. The masked-token loss is

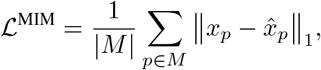

which encourages learning local structure and longer-range context without labels.

### G. Fine-Tuning Strategies

We compare four adaptation strategies for pre-trained encoders while keeping the training protocol fixed. Full fine-tuning updates all parameters end to end. Layer-wise unfreezing starts with task-specific layers and progressively unfreezes earlier blocks after warm-up [32]. LoRA freezes the original weights and adds trainable low-rank updates: convolutional LoRA for selected `Conv2d` layers in CNN backbones [33]. BNBitFit, which combines BitFit with batch-normalization affine tuning, updates only bias terms and batch-normalization affine parameters for lightweight distributional adjustment [34]. These strategies span full adaptation to highly restricted updates, allowing us to test whether parameter-efficient tuning is sufficient for dense prediction in small mammogram datasets.

### H. Loss Functions

We study losses targeting pixel-wise probability estimation, region overlap under class imbalance, boundary quality, and percent-density estimation. Standard losses include binary cross-entropy (BCE), Dice, IoU (soft Jaccard), Tversky, Focal, BCE+Dice, and Focal Tversky. BCE supports probabilistic calibration but can underweight rare foreground pixels. Dice, IoU, and Tversky optimize overlap under foreground–background imbalance, with Tversky controlling false positives and false negatives. Focal variants emphasize hard pixels but can be hyperparameter-sensitive in small datasets.

We also evaluate a density-calibrated hybrid objective that aligns mask learning with percent-density estimation. Motivated by bias in density estimates from predicted masks, it combines region-level, density-level, and boundary-level terms:

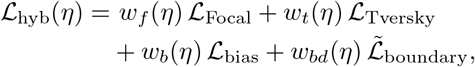

where *η* ∈ [0, 1] denotes normalized training progress. ℒ_Focal_ and ℒ_Tversky_ provide region-overlap supervision under class imbalance. ℒ_bias_ penalizes the absolute difference between predicted and ground-truth percent density. 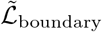 is a log-compressed boundary discrepancy term that encourages contour agreement while keeping the boundary penalty comparable to the other components.

The density-bias term applies the definition in Section II-B with predicted probabilities *p*_*i*_ in place of *ŷ*_*i*_, so that it is differentiable:

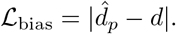

This tests whether directly penalizing density error improves percent-density calibration beyond overlap-based losses.

## III. Experiments

### A. Datasets and Data Preparation

We use the Mammogram Density Assessment (MDA) dataset [11], derived from VinDr-Mammo [35]. VinDr-Mammo contains 20,000 retrospectively collected mammo-grams from two primary hospitals in Hanoi, Vietnam. The released pixel-level density subset includes 596 mammograms with expert breast-area and dense-tissue annotations.

We split the labeled data into 405 training images (68%), 71 validation images (12%), and 120 test images (20%), with patient-level separation across L/R and CC/MLO views to avoid leakage. All inputs are resized to 512 × 512 pixels. Unless otherwise stated, single-channel mammo-grams are replicated to three channels and standardized using ImageNet statistics (mean [0.485, 0.456, 0.406], std [0.229, 0.224, 0.225]). Masks are resized with nearest-neighbor interpolation, and image intensities are normalized to [0, 1] per image before standardization.

### B. Pre-training Dataset and Setup

We use VinDr-Mammo [35] as the in-domain unlabeled corpus for SSL pre-training. It contains 5,000 four-view FFDM exams (L/R and CC/MLO; 20,000 images) with de-identified metadata. For SSL, we use only image pixels and discard all labels and exam-level annotations, ensuring no information leakage on ground-truth masks during testing or inference.

The default pre-training setting uses batch size 128, learning rate 1 × 10^−4^, 100 epochs, and mixed-precision training. SimCLR uses standard two-crop augmentation with temperature *τ* = 0.2. Multi-view SimCLR uses anatomical weights selected by grid search: 1.0 for same-side same-view pairs, 0.7 for same-side different-view pairs, 0.4 for different-side same-view pairs, and 0.2 for different-side different-view pairs. Barlow Twins uses off-diagonal penalty *λ* = 0.005. Masked image modeling uses mask ratio 0.6, patch size 16, an *l*_1_ reconstruction loss, and a minimum patch mean threshold of 0.02 to avoid masking pure background.

### C. Experimental Configuration

Supervised segmentation models are trained on the labeled MDA training subset. Unless otherwise noted, models use batch size 4, AdamW (weight decay 1 × 10^−4^), up to 50 epochs, and mixed-precision training. Full fine-tuning uses learning rate 1 × 10^−3^. Layer-wise unfreezing uses learning rate 3 × 10^−4^ with a 5-epoch warm-up and progressive unfreezing when applicable. LoRA uses rank *r* = 8, scaling *α* = 16, and encoder learning rate 1 × 10^−4^. For CNN backbones, LoRA is applied to selected `Conv2d` layers as low-rank residual updates. Backbone weights remain frozen except for adapted layers, while the segmentation head is always trainable. Configurations are selected sequentially— architecture, then SSL initialization, fine-tuning strategy, and loss—each step fixing the best choice from the previous one; this is a greedy search rather than a full factorial ablation.

The hybrid-loss weights are selected by grid search over a two-stage schedule. The final weights are set as follows:

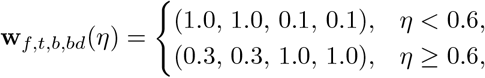

Early epochs emphasize mask learning through focal and Tversky terms; later epochs increase the density-bias and boundary terms to improve percent-density calibration and contour quality.

We report Dice similarity, Intersection over Union (IoU), and boundary F1 at a 2-pixel tolerance (BF1@2px). Dice and IoU are computed as

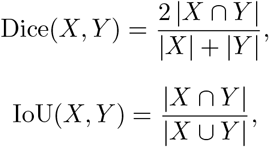

where *X* and *Y* denote the predicted and ground-truth masks. BF1@2px counts a predicted boundary pixel as correct if it lies within 2 pixels of the nearest ground-truth boundary. Logits are thresholded at 0.5, and the checkpoint with the highest validation Dice is evaluated once on the held-out test set. Reported values are means over test images from a single training run. Percent-density metrics follow Section II-B.

CNN backbones use a U-Net-style decoder, whereas U-Net-family models retain their original encoder–decoder architectures. ViT and DINOv3 backbones use a lightweight convolutional decoder with four upsampling blocks and skip connections from intermediate transformer features. MedSAM is evaluated separately with box-only, box + breast-region, and CNN-prediction prompts.

## IV. Results

### A. Classical and CNN/U-Net Standalone Baselines

We compare the classical baseline with standalone CNN and U-Net-family models under a unified training protocol. Unless otherwise specified, all deep models in Table I use ImageNet initialization, full fine-tuning, and BCE loss; later ablations use their stated SSL and fine-tuning settings. Results are summarized in Table I.

**TABLE I:** Standalone classical, CNN, and U-Net comparison for dense tissue segmentation on the held-out test set.

| **Group** | **Model** | **Dice** | **IoU** | **BF1@2px** |
|---|---|---|---|---|
| **Classical** | Adaptive FCM + SVM | 0.530 | 0.399 | 0.087 |
| **CNN** | VGG19 | 0.746 | 0.628 | 0.710 |
|  | ResNet50 | 0.781 | 0.666 | 0.786 |
|  | Xception | 0.798 | 0.690 | 0.776 |
|  | EfficientNet | **0.818** | **0.717** | **0.819** |
| **U-Net** | Vanilla U-Net | 0.692 | 0.566 | 0.650 |
|  | ResUNet | 0.775 | 0.659 | 0.773 |
|  | nnUNet | **0.790** | **0.692** | **0.786** |

CNN encoder–decoder models achieve the strongest standalone performance. EfficientNet performs best, with Dice 0.818, IoU 0.717, and BF1@2px 0.819. Xception and ResNet50 also perform well, highlighting the benefit of convolutional inductive bias under limited supervision. Within the U-Net family, nnUNet and ResUNet narrow the gap, suggesting that strong encoder–decoder architectures remain effective for small medical segmentation datasets. The adaptive fuzzy c-means + SVM baseline performs worst, with Dice 0.530, IoU 0.399, and BF1@2px 0.087, indicating that intensity-based methods are insufficient for robust dense-tissue segmentation in this low-contrast setting.

### B. Transformer and Prompt-Based MedSAM Analysis

We evaluate ViT and DINOv3 as standalone transformer baselines and MedSAM under separate prompt settings, since SAM-style models are prompt-dependent. Results are shown in Table II.

**TABLE II:** Transformer and prompt-based MedSAM results.

| **Method** | **Prompt** | **Dice** | **IoU** | **BF1@2px** |
|---|---|---|---|---|
| ViT | N/A | 0.684 | 0.549 | 0.090 |
| DINOv3 | N/A | 0.564 | 0.425 | 0.047 |
| MedSAM | Box-only | 0.511 | 0.371 | 0.096 |
| MedSAM | Box + breast region | 0.529 | 0.384 | 0.099 |
| MedSAM | CNN prediction | **0.794** | **0.680** | **0.250** |

ViT improves over adaptive fuzzy c-means + SVM in region overlap, with Dice 0.684, IoU 0.549, and BF1@2px 0.090, but remains below the CNN and U-Net-family models in Table I. DINOv3 performs worse than ViT, with Dice 0.564, IoU 0.425, and BF1@2px 0.047. Both transformer models have weak boundary scores, suggesting that pre-trained transformer features do not transfer directly to this task and that densetissue boundaries benefit from convolutional inductive bias.

MedSAM performance depends strongly on the prompt. Box-only and box + breast-region prompts yield low Dice scores of 0.511 and 0.529, respectively. Using the best CNN prediction as a dense prompt improves MedSAM to Dice 0.794, IoU 0.680, and BF1@2px 0.250, but still does not exceed the best standalone CNN configuration, including EfficientNet with BCE loss (0.818) or with the density-calibrated hybrid loss (0.837; Section IV-E). These results indicate that, even with a fine-tuned mask decoder, MedSAM does not improve over the CNN prediction it receives. One contributing factor is the decoder trained with CNN predictions on the training images, which are more accurate than the prediction at test time.

### C. Impact of Self-Supervised Pre-training

We test whether image-only SSL pre-training on VinDr-Mammo improves downstream dense-tissue segmentation. For the three strongest CNN/U-Net backbones, we evaluate masked image modeling (MIM), SimCLR, multi-view SimCLR, and Barlow Twins, followed by the same supervised fine-tuning protocol. Results are shown in Table III.

**TABLE III:**
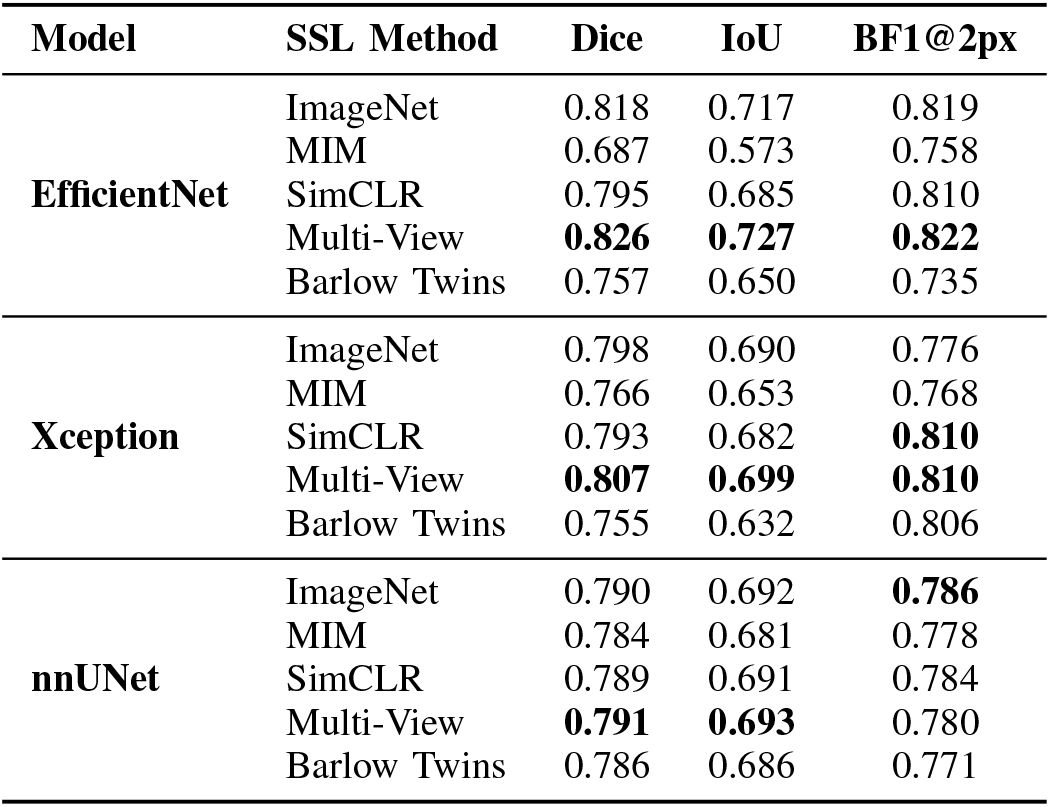
Effect of self-supervised pre-training on dense tissue segmentation for EfficientNet, Xception, and nnUNet.

SSL effects depend on both backbone and objective. For EfficientNet, MIM and Barlow Twins reduce performance, SimCLR remains below the ImageNet baseline, and only the multi-view objective improves results, reaching Dice 0.826, IoU 0.727, and BF1@2px 0.822. For Xception, the multi-view objective gives the best Dice and IoU, while SimCLR matches its BF1@2px. For nnUNet, all SSL variants remain close to baseline. Thus, SSL is useful mainly when the pretext task matches the small-label, multi-view mammography setting.

### D. Impact of Fine-Tuning Strategy

Using multi-view SSL initialization for EfficientNet, Xception, and nnUNet, we compare full fine-tuning, layer-wise unfreezing, LoRA, and BNBitFit. Results are shown in Table IV.

**TABLE IV:**
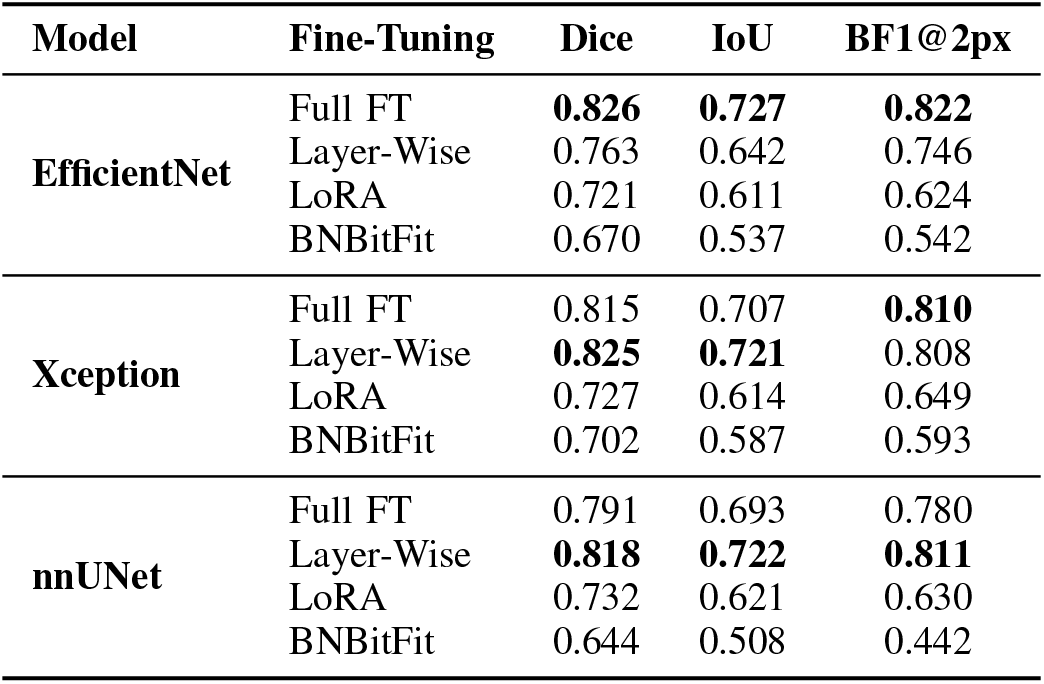
evaluation framework. A fixed bEffect of fine-tuning strategies for three backbones using multi-view SSL initialization.

| **Model** | **Fine-Tuning** | **Dice** | **IoU** | **BF1@2px** |
|---|---|---|---|---|
| **EfficientNet** | Full FT | **0.826** | **0.727** | **0.822** |
|  | Layer-Wise | 0.763 | 0.642 | 0.746 |
|  | LoRA | 0.721 | 0.611 | 0.624 |
|  | BNBitFit | 0.670 | 0.537 | 0.542 |
| **Xception** | Full FT | 0.815 | 0.707 | **0.810** |
|  | Layer-Wise | **0.825** | **0.721** | 0.808 |
|  | LoRA | 0.727 | 0.614 | 0.649 |
|  | BNBitFit | 0.702 | 0.587 | 0.593 |
| **nnUNet** | Full FT | 0.791 | 0.693 | 0.780 |
|  | Layer-Wise | **0.818** | **0.722** | **0.811** |
|  | LoRA | 0.732 | 0.621 | 0.630 |
|  | BNBitFit | 0.644 | 0.508 | 0.442 |

The best adaptation strategy depends on the backbone, but highly restricted methods consistently underperform. EfficientNet performs best with full fine-tuning (Dice 0.826, IoU 0.727), whereas Xception and nnUNet benefit more from layer-wise unfreezing. LoRA and BNBitFit drop across all three backbones, suggesting that their update capacity is insufficient for dense prediction in this small-data setting.

### E. Impact of Loss Function

We study segmentation losses while fixing architecture, SSL initialization, and fine-tuning strategy. For each of the three strongest backbones, we start from its best SSL and fine-tuning configuration from the preceding ablations and vary only the loss function. Results are reported in Table V.

Most standard losses perform similarly once a region-aware formulation is used. For EfficientNet, the hybrid loss gives the highest Dice and BF1@2px, but the gain over Tversky is small: Dice increases from 0.834 to 0.837, while IoU remains 0.739. The hybrid objective also gives the best Xception results, but reduces nnUNet performance, suggesting that its density and boundary terms are not universally beneficial. We therefore interpret it mainly as a density-calibration loss rather than a consistently superior segmentation loss.

### F. Percent-Density Calibration

Because percent density is clinically meaningful for breast cancer risk analysis, we test whether the density-calibrated hybrid loss improves percent-density calibration (Section II-B). Table VI compares the classical baseline, EfficientNet with Tversky loss, and EfficientNet with the hybrid loss, both with multi-view SSL and full fine-tuning.

The classical method has the largest density error (MAE 19.32, RMSE 24.97, bias 10.77) and weakest rank correlation (Spearman *ρ* = 0.279), consistent with its poor segmentation performance. Among EfficientNet models, the hybrid loss reduces test MAE from 14.81 to 11.78, RMSE from 23.13 to 19.54, and absolute bias from 6.05 to 4.71 percentage points, and increases Spearman correlation from 0.415 to 0.508. These results support the hybrid objective mainly as a percent-density calibration loss.

**TABLE V:** Effect of loss functions on dense tissue segmentation for the three strongest backbones.

| **Model** | **Loss Function** | **Dice** | **IoU** | **BF1@2px** |
|---|---|---|---|---|
| **EfficientNet** | BCE | 0.826 | 0.727 | 0.822 |
|  | Focal | 0.833 | 0.733 | 0.816 |
|  | Dice | 0.827 | 0.729 | 0.813 |
|  | IoU | 0.828 | 0.729 | 0.828 |
|  | Tversky | 0.834 | **0.739** | 0.824 |
|  | BCE+Dice | 0.826 | 0.727 | 0.822 |
|  | Focal Tversky | 0.815 | 0.708 | 0.805 |
|  | Hybrid | **0.837** | **0.739** | **0.830** |
| **Xception** | BCE | 0.825 | 0.721 | 0.808 |
|  | Focal | 0.825 | 0.722 | 0.811 |
|  | Dice | 0.797 | 0.691 | 0.779 |
|  | IoU | 0.808 | 0.701 | 0.780 |
|  | Tversky | 0.822 | 0.717 | 0.811 |
|  | BCE+Dice | 0.822 | 0.719 | 0.818 |
|  | Focal Tversky | 0.816 | 0.710 | 0.811 |
|  | Hybrid | **0.828** | **0.730** | **0.819** |
| **nnUNet** | BCE | 0.818 | 0.722 | 0.811 |
|  | Focal | 0.817 | 0.723 | 0.816 |
|  | Dice | 0.819 | 0.722 | 0.813 |
|  | IoU | **0.825** | **0.733** | **0.828** |
|  | Tversky | 0.820 | 0.726 | 0.824 |
|  | BCE+Dice | 0.820 | 0.726 | 0.822 |
|  | Focal Tversky | 0.819 | 0.725 | 0.805 |
|  | Hybrid | 0.804 | 0.701 | 0.780 |

**TABLE VI:** Percent-density estimation performance. Hybrid loss improves density calibration rather than mask overlap.

| **Model** | **Split** | **MAE** | **RMSE** | **Bias** | **Spearman** *ρ* |
|---|---|---|---|---|---|
| Adaptive FCM + SVM | Test | 19.32 | 24.97 | 10.77 | 0.279 |
| EfficientNet + Tversky | Train | 11.62 | 17.47 | −3.22 | 0.552 |
|  | Test | 14.81 | 23.13 | −6.05 | 0.415 |
| EfficientNet + Hybrid | Train | **9.15** | **13.12** | −1.67 | **0.660** |
|  | Test | **11.78** | **19.54** | −4.71 | **0.508** |

Figure 2 shows representative success and failure cases. Accurate predictions have high overlap and well-calibrated percent density. Failures mainly involve undersegmentation in extremely dense breasts, where fibroglandular tissue merges with posterior structures or residual tissue near the pectoral boundary, and false positives in low-density breasts, where vessel-like or noisy structures are mistaken for dense tissue.

**Fig 2:**
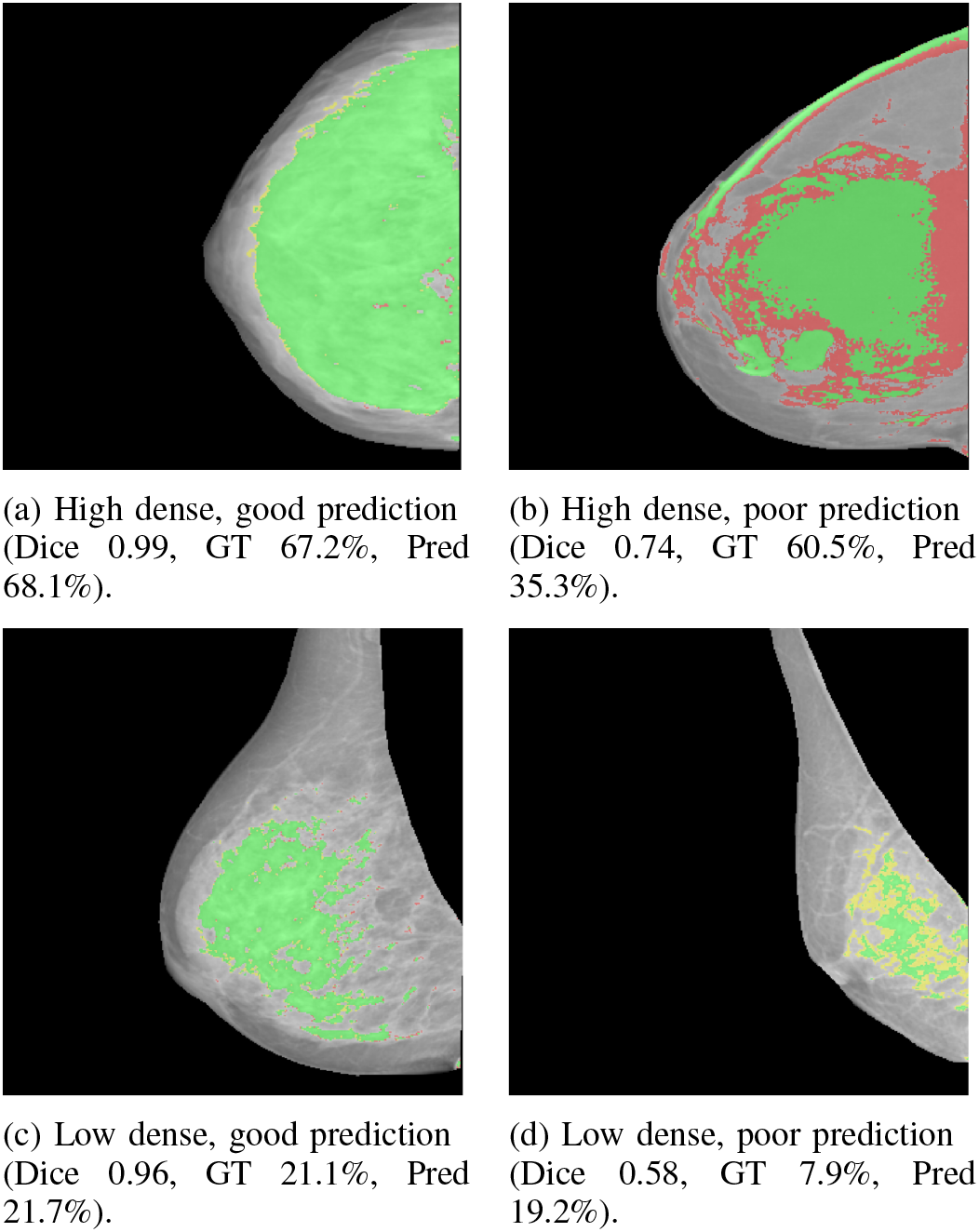
Segementation results for test samples using Efficient-Net with multi-view SSL, hybrid loss, and full fine-tuning. Green: true positives (TP), red: false positives (FP), yellow: false negatives (FN).

### G. External Clinical-Density Correlation Analysis

To assess clinical-density consistency beyond pixel-level segmentation, we correlate the final model’s percent-density estimates with clinical BI-RADS density categories across datasets. The final model is EfficientNet with multi-view SSL initialization, density-calibrated hybrid loss, and full fine-tuning. It is trained only on the MDA training split and evaluated on the held-out VinDr test set, InBreast [36], and MIAS [37].

InBreast and MIAS lack dense-tissue annotations compatible with our pixel-level protocol, so we use them for external clinical-density correlation rather than mask-overlap validation. For each image, we aggregate the predicted mask into a continuous percent-density estimate and measure its monotonic association with the available clinical density categories. The final model reaches Spearman correlations of 0.660 on the in-domain VinDr test set, 0.540 on InBreast, and 0.819 on MIAS, suggesting that predicted density scores preserve clinically meaningful ordering across datasets. Direct cross-domain Dice, IoU, and boundary evaluation remains future work pending compatible expert dense-tissue masks.

### H. Computational Cost and Practical Trade-offs

We measure fine-tuning and SSL pre-training time on a single NVIDIA H100 GPU. Fine-tuning takes approximately 0.95 hours for EfficientNet-B0, 1.00 hour for Xception, and 1.25 hours for ViT-Base. SSL pre-training adds 6–21 GPU hours depending on the backbone and objective, yet several SSL variants fail to improve over ImageNet initialization for EfficientNet, and ViT remains less accurate than CNN-based models despite higher fine-tuning cost. MedSAM refinement also adds approximately 1 GPU hour of mask-decoder fine-tuning and additional inference after CNN prediction without outperforming the standalone CNN model. The most practical configuration is therefore a compact CNN encoder–decoder model with task-aligned training and density calibration.

## V. Discussion and Conclusion

This study provides several practical lessons for dense breast tissue segmentation under limited pixel-level annotation. First, *task-appropriate inductive bias may matter more than model scale or recency in this setting*. Compact CNN encoder–decoders achieved stronger performance than ViT- and DINOv3-based alternatives, suggesting that local spatial modeling remains valuable for diffuse, low-contrast tissue segmentation. Similarly, LoRA and BNBitFit underperformed full or layer-wise fine-tuning, suggesting that dense prediction may require broader representation adaptation than highly restricted parameter-efficient methods.

Second, *self-supervised pretraining was most useful when it reflected mammography-specific structure*. Generic MIM, SimCLR, and Barlow Twins provided little or inconsistent benefit over ImageNet initialization, whereas multi-view SimCLR yielded the strongest SSL results for EfficientNet and Xception. This suggests that exploiting domain-specific relationships, such as laterality and complementary views, may be more useful than applying generic SSL objectives to additional unlabeled images.

Third, *the MedSAM results highlight the importance of prompt informativeness and task geometry*. Box-based prompts performed poorly, whereas a dense CNN-generated prompt substantially improved MedSAM; however, the refined output still did not exceed the CNN prediction used as the prompt. For diffuse tissue segmentation, simple prompts may provide less task-relevant information than they do for compact-object segmentation. Foundation models should therefore be evaluated by the incremental value they provide beyond the model or information source used to generate the prompt.

Finally, *segmentation overlap and downstream quantitative accuracy are related but distinct objectives*. The densitycalibrated hybrid loss provided only modest improvement in Dice but more clearly reduced percent-density error and bias, suggesting that optimizing overlap alone may not optimize clinically meaningful measurements. More broadly, additional model complexity should be justified by measurable gains relative to its computational and implementation cost.

This study is limited by the small labeled dataset and singlerun evaluation, so small performance differences should be interpreted cautiously. External validation is also limited by the lack of compatible pixel-level annotations in InBreast and MIAS. Future work should evaluate larger multi-center cohorts, repeated runs with statistical testing, and broader task-specific adaptation of foundation models.

## Data Availability

The datasets used in this study are publicly available, including VinDr-Mammo, InBreast, and MIAS. These datasets can be accessed through their respective official repositories subject to registration or data use agreements. All other data generated or analyzed during this study are included in the manuscript.

## Acknowledgment

Zhijun Yin and Xingyi Guo are supported by the National Cancer Institute (NCI) of the National Institutes of Health (NIH) under grant R01CA297582.

